# Differential expression patterns of sIgA in Human Breast milk (HBM) is dependent on birth outcome

**DOI:** 10.1101/2025.02.19.25322535

**Authors:** Gareth A Nye, Magdalena J Rzepecka, Christopher E Jones, Kate Harrison

**Affiliations:** School of Science, Engineering and Environment, University of Salford, Manchester, M5 4WT; Chester Medical School, University of Chester, Chester, CH1 4BJ

## Abstract

Breast milk is key for the development of newborns, particularly their immune systems and gut microbiota. In times of neonatal care, newborns are often supplemented with donor breast milk for a range of practical and medical reasons. However, we do not currently understand whether specific breast milk samples may be better suited at boosting the immune system. One of the most influential immune components is sIgA immunoglobulin.

**Methods:** Donor human breast milk samples provided by the North West Human Milk Bank were analysed for levels of sIgA using Abnova sIgA (Human) ELISA Kit according to the manufacturer’s instructions and analysed via statistical software packages based on the anonymised maternal characteristics.

**Results:** sIgA levels were significantly increased in breast milk samples following preterm and stillbirth outcomes compared with term and live deliveries. In preterm deliveries, sIgA levels remained significantly higher in breast milk for a longer postnatal period when compared with term deliveries. There was no significant changes in sIgA levels with antibiotic use.

**Conclusion:** The results presented in this study suggest that human breast milk is tailored to the baby from an immunological perspective. Higher levels of sIgA in breast milk being seen in pregnancies which did not end in a healthy baby i.e. pregnancies ending in preterm delivery or stillbirth would suggest there is an internal mechanism within the mother to provide additional support to a baby which is failing to grow successfully. This may open up new avenues to select donor samples specifically to assist in babies which are born premature to improve their immune systems.

**Key Messages:**

- **Research has long shown that human breast milk is best for the development of newborn immune systems and gut microbiota**
- **In neonatal settings, donor breast milk is used to supplement in times of need**
- **This study shows for the first time that immunological factors can be altered based on fetal outcome measures**
- **This may inform the provision of donor breast milk in neonatal feeding to ensure maximum immune development in preterm babies**.

## Introduction

Breast milk is a complex fluid that provides optimal nutrition and immunological protection for infants. It is the recommended feeding method for all infants, particularly for those born prematurely (1).

Breast milk should provide essential nutrients for growth and development during the critical first few months of life. However the nutritional composition of breast milk can vary widely between mothers and even within the same mother over time. Additionally it can be affected by a range of factors, including maternal diet, genetics, lactation stage, and environmental factors (2).

Despite the high variation between samples, we have strong evidence to suggest that breast milk from term pregnancies typically contains higher levels of fat and lactose, while breast milk from preterm pregnancies contains higher levels of protein and minerals (3). Preterm breast milk is also higher in calories and growth factors that are essential for the development of preterm infants (4, 5).

In addition to its nutritional content, breast milk also contains a wide range of immunological factors, including antibodies, cytokines, and other immune cells, which provide passive immunity to the infant (6, 7). These factors play a critical role in protecting the infant from infections and other diseases and is of particular interest for those babies born prematurely. However, the immunological content of breast milk can be hugely impacted by maternal circumstances including infection and subsequent treatment during the pregnancy (1, 8). Even the use of antibiotics during pregnancy has a potential impact on the immune power of human breast milk. It has also been well documented that use of cannabinoids during or shortly after pregnancy can dramatically lower the levels of immune components potentially leading to increased risk of infection in the newborn. (9)

Understanding the factors that influence the immunological content of breast milk is essential for optimizing infant health and well-being.

### Fundamentals and current medical usage of human breast milk

Breast milk is produced in the mammary glands of the breast in response to the hormone prolactin, which is secreted by the pituitary gland in the brain. The production and release of breast milk is controlled by the hormone oxytocin, which is released during breastfeeding and causes the milk to be released from the mammary glands into the ducts and out of the nipple (10).

Breast milk is typically divided into two main types: colostrum and mature milk. Colostrum is the first milk produced by the mother in the first few days after giving birth. It is a thick, yellowish fluid rich in immunological factors, such as antibodies, cytokines, and growth factors, which provide passive immunity and help to support the infant’s immune system (11). Colostrum also has a laxative effect, which helps to expel meconium and prevent jaundice (12).

After the first few days, the mother’s milk transitions to mature milk, which is thinner and whiter in colour than colostrum. Mature milk is composed of two main types of milk: foremilk and hindmilk. Foremilk is released at the beginning of a feeding, while hindmilk is released towards the end of a feeding. Foremilk is higher in lactose and lower in fat, while hindmilk is higher in fat and lower in lactose. The combination of foremilk and hindmilk provides the infant with a balance of nutrients and energy (13).

Donor human breast milk is used as a source of nutrition for babies under neonatal conditions where practical breast feeding is not possible. It is much preferred over formula in theses situations due to the infants underdeveloped immune system in addition to human breast milk being easier to digest and process. However a new wave of medical research into the use of human breast milk has bloomed with the major uses summarised in table 1.

**Table 1.** Summary of the current usage of donor human breast milk in medicine.

| Usage | Rationale |
| --- | --- |
| Neonatal medicine | Human breast milk is the preferred source of nutrition for premature infants, who are at increased risk of developing complications due to their underdeveloped immune systems and digestive systems. As well as the range of immunological and anti-inflammatory factors present that can help to protect premature infants from infections, human milk is easier for premature infants to digest compared to formula, which can help to reduce the risk of gastrointestinal issues. (14) |
| Gastrointestinal disorders | Breast milk has been shown to have beneficial effects on a range of gastrointestinal disorders, including inflammatory bowel disease (IBD), necrotizing enterocolitis (NEC), and gastroesophageal reflux disease (GERD). (15) |
| Wound healing | Human breast milk contains a range of growth factors and immune factors that may promote wound healing. Studies have shown that applying breast milk topically to skin wounds can help to speed up the healing process and reduce the risk of infection (16). |
| Cancer treatment | Researchers are currently investigating the potential use of breast milk components in cancer treatment, particularly in the development of new immunotherapies (17) |
| Microbiome modulation | Breast milk contains a range of prebiotic and probiotic factors that can help to modulate the infant's gut microbiome, which has been shown to play a key role in immune system development and overall health. (18) |

### Targeting human breast milk as a source of immunological factors

All infants are born with an immature and naïve immune system, in particular lacking immunoglobulin A, and it is well established that immunity can be passed from mother to an infant through maternal breast milk (19). This is particularly evident from the colostrum, which contains most of the immunological protection for the infant during the transition from intrauterine to extrauterine life (20). When compared with infants who are exclusively formula fed, breast fed infants have a reduced risk of developing significant gastrointestinal diseases (21) such as necrotizing enterocolitis (NEC) (22) and gastroenteritis (23) as well as reducing the likelihood of severe respiratory illness (24) and urinary tract infections (25)

### Specific immunological factors in HBM

The immunological factors contained within the maternal milk, such as immunoglobulins, live microbes, metabolites, immune cells, and cytokines are key in the early development of neonatal gut microbiota (26, 27).

Immunoglobulins are one of the most important anti-microbial bioactive factors within maternal milk. Maternal milk contains mainly Immunoglobulin A (IgA), M (IgM), and G (IgG) antibodies, where IgA constitutes approximately 90-95%, IgM to 2-5%, and IgG to <1% of all antibodies (28, 29).

Secretory IgAs are a part of the adaptive immunity provided by the breastmilk which is mediated by the maternal mucosal-associated lymphoid tissue (MALT) pathway, responsible for the stimulation of antigen-specific T and B lymphocytes (28, 30). In addition, sIgA is primarily involved in immune exclusion whereby mucosal surface protection is achieved by a direct cross-linking of environmental microorganisms or macromolecules to prevent contact with epithelial cell’s surface, which results in their elimination by peristalsis or mucociliary movement (31). The sIgA has also been noted to provide a balance in the mucosal surface which then ensures the survival of the mucosal florals (31)

IgA deficiency is a relatively common immunodeficiency disorder, which can lead to recurrent infections, particularly of the respiratory and gastrointestinal tracts which may stem from a lack of breast milk from birth (32).

To this date, human breast milk has not been adequately screened for levels of sIgA which may prove to be revolutionary in the priming of the immune system in premature infants who classically struggle with increased lifelong risk of infections.

This study aims to analyse the sIgA levels of human breast milk and identify particular subsets of donors who may improve the outcomes of premature babies in neonatal units. In essence, the results of this study can serve as a guide to the human breast milk banks globally to ensure that breast milk rich in specific immunological properties is received to match the intended needs.

## Methods

### Sample Collection

All human breast milk samples used in this study were obtained from the North-West Human Milk Bank, Chester, United Kingdom. The Milk Bank at Chester is the largest NHS milk bank in the England and provides donor milk to over 70 hospital neonatal units.

Milk goes through a filtration process following by microbiology screen and pasteurisation procedure before being frozen for later use.

One hundred and seventeen breast milk samples that could not be used for donation but were designated for research were collected from the milk bank along with anonymised information from the corresponding consent forms.

### Sample Preparation

Each breast milk sample was fully defrosted before being centrifuged for 15 minutes at 500g and 4°C. The supernatant was centrifuged for 15 minutes at 3000g and 4°C which was then diluted 1:500 in Phosphate Buffered Saline (PBS) for analysis.

### sIgA Content Analysis

The concentration of sIgA in human breast milk samples was measured using Abnova sIgA (Human) ELISA Kit according to the manufacturer’s instructions. Briefly, 195µL of red EIA buffer was pipetted into the microwells allocated for breast milk samples in duplicates followed by 10µL of the diluted samples. The contents in the microwells were carefully mixed and the plate was covered with adhesive tape before incubation for 90 minutes at 37°C. The plate was washed three times with the washing solution provided followed by the application of 100µL of the Conjugate. The plate was incubated for a further 30 minutes at 37°C. Following an addition wash period, 100µL of substrate solution was dispensed into the microwells and incubated for 15 minutes at 25°C. 100µL of stop solution was added to the microwells, and the absorbance was immediately measured at 450nm using a microplate reader.

### Statistical Analysis

GraphPad Prism 8.0 (GraphPad. Software, San Diego, CA, USA) was employed for the statistical analysis. The Mann–Whitney U test was applied to determine intergroup differences with Kruskal-Wallis test for additional measures. A p-value of less than 0.05 was considered significant.

## Results

Table 2 shows detailed nutritional values from the collected human breast milk samples. There is no significant difference in any of the recorded values when comparing pregnancies ending in caesarean section or vaginal delivery. During the 6 months following birth, we see significant decreases in protein levels only from month 3 onwards. The remaining nutritional measures stay consistent throughout the 6-month postnatal period.

**Table 2.** Nutritional values from the collected human breast milk samples with relation to delivery method and listed as values separated by the time since delivery. Asterixs refers to significant values compared with month 1 (p=<0.05). SD (standard deviation), CS (Caesarean Section), VD (Vaginal Delivery).

|  |  |  | Fat | Protein | Carbohydrate | Solids | Calories |
| --- | --- | --- | --- | --- | --- | --- | --- |
| Method of delivery | CS | Mean | 3.62 | 1.02 | 8.55 | 13.39 | 72.09 |
|  |  | SD | 1.00 | 0.22 | 0.27 | 1.18 | 9.76 |
|  | VD | Mean | 3.54 | 1.03 | 8.42 | 13.16 | 70.70 |
|  |  | SD | 0.92 | 0.26 | 0.42 | 1.09 | 9.08 |
| Time since delivery (Months) | 1 | Mean | 3.94 | 1.40 | 8.53 | 14.07 | 76.78 |
|  |  | SD | 0.66 | 0.24 | 0.23 | 0.72 | 6.22 |
|  | 2 | Mean | 3.19 | 1.06 | 8.53 | 12.93 | 67.88 |
|  |  | SD | 0.74 | 0.12 | 0.13 | 0.85 | 7.64 |
|  | 3 | Mean | 3.46 | **1.02 \*** | 8.64 | 13.30 | 71.00 |
|  |  | SD | 1.36 | 0.19 | 0.24 | 1.64 | 13.53 |
|  | 4 | Mean | 3.64 | **0.87 \*** | 8.56 | 13.30 | 71.86 |
|  |  | SD | 0.89 | 0.08 | 0.23 | 1.09 | 9.01 |
|  | 5 | Mean | 3.54 | **0.90 \*** | 8.52 | 13.17 | 70.56 |
|  |  | SD | 1.28 | 0.13 | 0.35 | 1.45 | 12.22 |
|  | 6 | Mean | 3.60 | **0.89 \*** | 8.23 | 12.87 | 69.82 |
|  |  | SD | 0.92 | 0.05 | 0.59 | 0.92 | 8.00 |

Upon investigating the levels of sIgA in these samples we have found significant differences in pregnancies ending in a live birth compared with a still born in addition to significant differences in whether the pregnancy ended at term or before the due date. We see higher mean values of sIgA in milk samples from both preterm and still born pregnancies compared with live, term deliveries. There was no significant difference relating to the mode of delivery (Table 3).

**Table 3.**
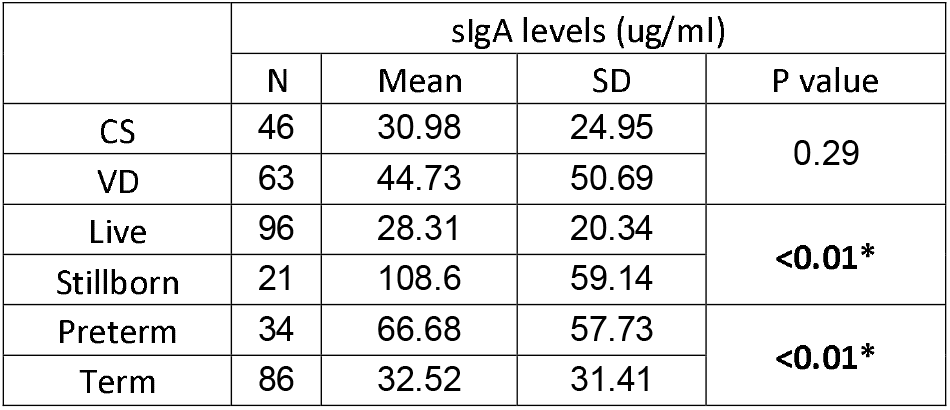
sIgA levels expressed as ug/ml with mean, standard deviation and P value. Comparisons are made in pairs based on birth outcome. Asterixs refers to significant values based on p value. SD (standard deviation), CS (Caesarean Section), VD (Vaginal Delivery).

|  | sIgA levels (ug/ml) |
| --- | --- |

|  | sIgA levels (ug/ml) |  |  |  |
| --- | --- | --- | --- | --- |
|  | N | Mean | SD | P value |
| CS | 46 | 30.98 | 24.95 | 0.29 |
| VD | 63 | 44.73 | 50.69 |  |
| Live | 96 | 28.31 | 20.34 | <0.01* |
| Stillborn | 21 | 108.6 | 59.14 |  |
| Preterm | 34 | 66.68 | 57.73 | <0.01* |
| Term | 86 | 32.52 | 31.41 |  |

[utble]

In addition, we investigated the change in sIgA levels from breast milk samples over the 6 month postnatal period in both preterm and term pregnancies. In both groups, the level of sIgA was significantly higher in milk samples extracted at 3 months post delivery when compared with samples extracted 1 month following delivery.

Following month 3, we see a differential pattern of sIgA levels between term and preterm. In term pregnancy samples we see a significant decrease in sIgA levels with reported levels in samples extracted at month 4 and 5 being significantly lower than month 1 samples. We do not see this significant decline in samples taken from preterm pregnancies which is compounded by a significant difference between the two groups in samples extracted at 4 months (Table 4).

**Table 4.** sIgA levels from samples of breast milk from month 1 to month 6 following birth expressed as ug/ml with mean, standard deviation and P value. Comparisons are made between month 1 and additional time points and between term and preterm births. Asterixs refers to significant values based on p value. SD (standard deviation), CS (Caesarean Section), VD (Vaginal Delivery).

|  | Preterm |  |  |  | Term |  |  |  | P value compared with preterm |
| --- | --- | --- | --- | --- | --- | --- | --- | --- | --- |
|  | N | Mean | SD | P value compared with month 1 | N | Mean | SD | P value compared with month 1 |  |
| Month 1 | 7 | 43.71 | 24.67 |  | 31 | 32.29 | 21.79 |  | 0.23 |
| Month 2 | 2 | 65.00 | 32.53 | 0.34 | 18 | 41.28 | 32.54 | 0.25 | 0.34 |
| Month 3 | 12 | 113.33 | 65.99 | <0.01 * | 4 | 102.50 | 79.62 | <0.01 * | 0.79 |
| Month 4 | 4 | 73.00 | 46.02 | 0.19 | 16 | 20.44 | 16.49 | 0.06 * | <0.01 * |
| Month 5 | 4 | 21.50 | 12.15 | 0.13 | 6 | 12.83 | 4.45 | 0.04 * | 0.14 |
| Month 6 | 2 | 13.50 | 7.78 | 0.15 | 11 | 21.73 | 12.23 | 0.14 | 0.39 |

A comparison of sIgA levels in human breast milk from mothers who used antibiotics and those who did not was also conducted. The data included 9 gestation matched samples which showed a mean sIgA level in the antibiotic group of 33.22, and in the non-antibiotic group, a mean sIgA level of 31.22, which was not significant.

## Discussion

The results from this study provide valuable insights into the nutritional composition of human breast milk and its variations under different maternal and postnatal conditions. In addition, we have highlighted significant differences in the immunological profile of milk samples based on pregnancy outcomes.

### Nutritional Values and Mode of Delivery

The finding that there is no significant difference in the nutritional values of breast milk between cesarean and vaginal deliveries suggests that the mode of delivery does not impact the overall nutritional profile of breast milk. This aligns with previous research indicating the macronutrient content of breast milk is stable across different delivery methods (33, 34). This finding is crucial for healthcare practitioners and expectant mothers, as it indicates that Caesarean section deliveries do not compromise the nutritional quality of breast milk. The lack of significant differences may also reflect robust biological mechanisms that ensure the infant receives adequate nutrition regardless of the mode of delivery. More work needs to be conducted to establish whether the underlying causes for the Caesarean section (e.g. fetal distress, maternal complications) can influence the breast milk (35).

### Postnatal Changes in Protein Levels

The significant decrease in protein levels beginning from the third month postpartum aligns with existing literature indicating that protein concentrations in breast milk typically reduce as lactation progresses (36-38). This decrease may be attributed to the changing nutritional needs of the growing infant and the gradual adaptation of breast milk composition. This trend underscores the dynamic nature of breast milk composition and highlights the importance of continuous monitoring of protein intake in infants to ensure optimal growth and development (37). It also raises questions about the adequacy of protein intake for infants beyond the third month of lactation and whether supplemental feeding might be necessary, which may be of great importance in the feeding of infants in hospital settings who are almost wholly reliant on donated breast milk (39).

### sIgA Levels and Pregnancy Outcomes

The study’s investigation into sIgA (secretory Immunoglobulin A) levels in breast milk provided several noteworthy findings. sIgA is a crucial component of the immune system, playing a significant role in protecting mucosal surfaces from pathogens. It provides passive immunity to infants, helping to defend against infections until their own immune system matures (40).

### Differences Between Live Births and Stillbirths

The observation of significantly higher sIgA levels in breast milk from pregnancies that ended in stillbirth reveals intricate aspects of maternal immunological responses. A possible explanation for the elevated sIgA levels could be related to the heightened maternal immunological activation during pregnancies with complications leading to stillbirth. The maternal immune system might respond to stress, infection, or inflammation associated with adverse pregnancy outcomes by increasing the production of immunoglobulins, including sIgA. This response might be an adaptive mechanism aimed at providing the newborn, if born alive, with enhanced immune protection against potential pathogens.

Interestingly, maternal infections or inflammatory conditions during pregnancy, such as chorioamnionitis, have been shown to stimulate an increased secretion of sIgA in breast milk (41, 42). In the context of stillbirths, a similar underlying inflammatory process could lead to the upregulation of sIgA production. Further research is necessary to confirm this hypothesis and elucidate the precise pathways involved.

### Impact of Maternal Health and Nutrition

Maternal health and nutritional status are critical factors influencing breast milk composition. It is plausible that mothers who experienced stillbirths might have had underlying health conditions impacting immune function and breast milk qualities. For example, maternal conditions such as preeclampsia or gestational diabetes are known to affect immune responses and could potentially contribute to the observed differences in sIgA levels (43).

Additionally, the emotional and psychological stress associated with stillbirths might also play a role. Stress and emotional strain can affect immune function, potentially leading to alterations in breast milk composition. Chronic stress has been linked to reduced levels of certain immune components, including sIgA, as the body attempts to bolster its defences during perceived threats or challenges itself rather than transfer (44, 45)

The findings of higher sIgA levels in breast milk following stillbirths have important clinical implications. Understanding the underlying causes of these differences can aid in developing targeted interventions to support maternal and infant health. For instance, providing additional support and monitoring for mothers who experience stillbirths could help ensure optimal breastfeeding outcomes and the immunological protection of breast milk.

Future research should focus on comprehensive investigations into the factors contributing to elevated sIgA levels in cases of stillbirth. This includes examining maternal immune function, inflammatory markers, and stress responses during and after pregnancy. Additionally, longitudinal studies tracking the health outcomes of infants receiving varying sIgA levels could provide valuable insights into the long-term benefits and potential risks associated with altered breast milk composition.

### Differences Between Term and Preterm Deliveries

Higher sIgA levels were also observed in preterm deliveries compared to term deliveries. This could be attributed to the additional immunological protection needed for preterm infants, who are more vulnerable to infections due to their underdeveloped immune systems. The elevated sIgA levels in breast milk following preterm births might reflect the mother’s immune system’s effort to provide enhanced protection to the infant. This underscores the critical importance of breast milk for preterm infants and supports the practice of early and exclusive breastfeeding in this group.

### Patterns Over the Postnatal Period

Longitudinal data indicated that while sIgA levels peaked around the third month postpartum in both term and preterm pregnancies, the subsequent patterns diverged. In term pregnancies, sIgA levels experienced a sharp decrease after the third month, dropping significantly below the levels seen in the first month by the fourth and fifth months. This sharp decline might suggest that term infants gradually develop their own immune defences, reducing the need for high levels of maternal immunoglobulins in the breast milk.

Conversely, the decline in sIgA levels in preterm pregnancies was less pronounced, and a significant difference in sIgA levels between term and preterm samples was observed by the fourth month. This suggests that preterm infants continue to rely heavily on the immunoprotective benefits of breast milk beyond the third month, necessitating a prolonged period of higher sIgA delivery through breast milk. This pattern highlights the need for targeted breastfeeding support and potential supplementation strategies for preterm infants to ensure they receive adequate immunological protection during their critical early developmental stages.

These findings highlight that maternal and pregnancy-related factors significantly influence the immunological composition of breast milk. The elevated sIgA levels in preterm and stillborn pregnancies suggest a sophisticated maternal adaptive mechanism aimed at supporting neonatal immunity. However, the exact biological pathways and the influence of other potential confounders, such as maternal diet and health status, need further exploration. The differential decline rates of sIgA in term versus preterm samples also underscore the necessity of individualized breastfeeding guidance, particularly for mothers of preterm infants who may require extended periods of high sIgA levels to safeguard infant health. This may also be used by milk banks to ensure the most appropriate samples are being provided to high-risk babies in neonatal care, ensuring appropriate growth and immune system development.

Future research should focus on elucidating the mechanisms behind these observations and investigating the long-term health outcomes for infants exposed to varying levels of sIgA. This could include studies on how maternal health interventions might optimize sIgA concentrations in breast milk and the potential benefits of supplemental immunoglobulin strategies for at-risk infant populations.

### Impact of Antibiotics on sIgA Levels

The comparison of sIgA levels between mothers who used antibiotics during pregnancy and those who did not showed no significant difference, suggests that antibiotic use during pregnancy does not significantly alter the sIgA concentration in breast milk. This finding is reassuring for clinical management however, it also highlights the need for further studies to investigate the potential impacts of different types of antibiotics on breast milk composition and the infant’s developing immune system. This however does go against recent animal studies which suggest that prenatal antibiotic usage can significantly decrease the levels of sIgA in mouse breast milk (46). It does raise the question of maternal infection and when the antibiotics were used in the pregnancy period and even if any underlying infection could change the immune contents of the breastmilk (47)

## Conclusion

In summary, this study highlights the importance of understanding how maternal and postnatal factors influence the nutritional and immunological properties of breast milk. These findings have direct implications for clinical practice, breastfeeding guidance, and infant nutrition strategies, particularly for preterm infants who may require tailored support to harness the full immunological benefits of breast milk. Future research should continue to explore the underlying mechanisms driving these changes and assess the long-term health impacts on infants. Additionally, these results underscore the need for healthcare providers to closely monitor the nutritional status of both mothers and infants, and to provide individualized support to optimize breastfeeding outcomes

## Data Availability

All data produced in the present work are contained in the manuscript

